# Climate change worry and the association with future depression and anxiety: cross-national analysis of 11 European countries

**DOI:** 10.1101/2024.09.02.24312950

**Authors:** A. Collery, C.L. Niedzwiedz

## Abstract

**BACKGROUND:** Climate change is one of the biggest threats to global health and can affect people’s mental health directly and indirectly. This study focused on the implications of climate change worry for mental health.

**OBJECTIVE:** To determine the association between climate change worry and future risk of depression, anxiety and sleep disturbance across 11 European countries.

**METHODS:** The study included 5,155 participants in the European Social Survey-10 (2020-22), which assessed climate change worry and subsequent follow-up CROss-National Online Survey 2 (CRONOS-2) wave 4, which measured symptoms of depression and anxiety (2022). Overall relationships between climate change worry and risk of depression, anxiety and sleep disturbance were explored using logistic regression models adjusting for potential confounders, then stratified by country.

**FINDINGS:** Climate change worry was associated with increased risk of anxiety (OR: 1.38, 95% CI: 1.13-1.68), but not depression (OR: 1.10, 95% CI: 0.94-1.29) or sleep disturbance (OR: 1.08, 95% CI: 0.92-1.27), in pooled analyses across countries. When stratified by country, climate worry had differing associations with the outcomes, with the strongest relationships between climate worry and anxiety found in Slovenia and Italy.

**CONCLUSIONS:** Climate change worry may contribute to clinically significant anxiety in some countries, but less so for depression and sleep disturbance.

**CLINICAL IMPLICATIONS:** Strategies to cope with climate change anxiety are needed as it does not align with traditional mental health models. Further research is needed to design and evaluate interventions and policies to support mental health in the context of climate change.

**What is already known on this topic:** Climate anxiety is a growing concern as the implications of climate change become more evident. Few studies have assessed how climate worry is related to mental health problems and research in this area is limited by methodological concerns, such as non-representative samples and cross-sectional studies.

**What this study adds:** Levels of climate-related worry vary across Europe, with the highest levels observed in Portugal, Austria and Slovenia, and lowest in Sweden, Iceland and Italy. Overall, climate worry was associated with future risk of clinical levels of anxiety, but not depression or sleep problems. In stratified analyses, the strongest associations were found for anxiety in Slovenia and Italy.

**How this study might affect research, practice or policy:** The mental health implications of climate change should be considered in both health and climate policies. Further research is required to assess country and regional-level factors which may affect the relationship between climate worry and mental health and potential causal effects investigated.

## BACKGROUND

Climate change is a growing global health crisis primarily driven by combustion of fossil fuels which result in rapidly rising carbon dioxide emissions. This emergency poses significant threats to health through direct and indirect effects. Direct effects of climate change on health include consequences of extreme weather events, such as heatwaves, floods and droughts [1]. Conversely, indirect effects are described as threats to social, economic and environmental determinants of health [2], such as changes in geographical patterns of vector-borne diseases, food insecurity and threats to mental health [1]. An emerging phenomenon is the concept of ‘eco-anxiety’, defined as “people’s reactions of worry and anxiety in view of global climate change threats and concurrent environmental degradation” [1]. This concept focuses on the general awareness of climate change rather than a response to traumatic extreme weather events [3]. There are gaps in knowledge relating to the prevalence of eco-anxiety, and whether it is associated with specific mental health problems and symptoms [4]. Previous studies have suggested that eco-anxiety is linked to functional impairment, symptoms of depression and anxiety, post-traumatic stress disorder (PTSD) and insomnia [1]. However, research in this area generally lacks quality due to poor study designs. For instance, the majority of studies employ a cross-sectional design, therefore preventing causal inferences to be made. Studies have also begun to identify groups vulnerable to worry about climate change: women, people living in poorer countries and younger people [4], [5]. The role of other variables such as education, culture and region remains unknown [3].

There are many reasons for climate change worry. Awareness of climate change is extensive, with the media providing wide coverage on the issue and usually with a pessimistic narrative [3]. Climate change can lead to feelings of helplessness, fear and grief [2]. Many young people in particular feel frustrated and betrayed by governments who are failing to act against climate change, therefore putting their futures in jeopardy [6]. This is further driven by individual-level knowledge and education, which can empower people into climate action, but can also have the opposite effect and induce a sense of hopelessness and overwhelm [3].

Worry about climate change can have multiple consequences. It can lead to disempowerment – when feeling that efforts to reduce climate change are inadequate, as well as feelings of isolation among youth due to dismissal of peers and adults around them when expressing their worries. However, in some studies climate change worry has been linked to a green identity and adoption of pro-environmental behaviours [1]. Interestingly, the opposite has also been found in other studies, with climate change worry found to be associated with disengagement from pro-climate actions [1], potentially due to feeling overwhelmed at the extent of the situation [7]. Lastly, there are potential links between worry about climate change and negative mental health outcomes, including insomnia [1]. Mental disorders are a leading cause of disability, the burden of which is continuing to rise globally [8].

Therefore, if climate worry is related to adverse mental health outcomes, strategies are needed to channel it into positive action to reduce any potential mental health burden.

## OBJECTIVES

This study aimed to investigate whether worry about climate change was associated with risk of depression, anxiety and sleep disturbance across Europe. The objectives were three-fold: (i) to determine which countries have the highest levels of worry about climate change; (ii) to determine if high levels of climate change worry are associated with future risk of depression, anxiety and sleep disturbance; and (iii) to determine which countries had the strongest associations between climate change worry and risk of depression, anxiety and sleep disturbance.

## METHODS

### Study Design and Participants

This study utilised data from the 10^th^ European Social Survey (ESS) (collected during 18/09/2020 - 03/09/2022) [9], [10], and subsequent follow-up CROss-National Online Survey 2 (CRONOS-2) wave 4 (collected during 03/05/2022 - 22/12/2022)[11], allowing for a longitudinal study design. The data are openly accessible (upon registration) at https://www.europeansocialsurvey.org/. ESS10 is a cross-national survey conducted across 31 countries [12]. During this survey round data were collected by a face-to-face interview (via computer-assisted personal interview, web-based video interview, or paper/web self-completion methods). The target population was people aged 15 years and over resident within private households, regardless of their nationality, citizenship, language or legal status [12]. Individuals were selected by strict random probability methods, including simple, stratified and multistage methods. Further detailed information regarding the methodology of ESS10 is found elsewhere [12].

CRONOS-2 is a cross-national, probability-based input-harmonised web panel. It was conducted across 12 of the countries which took part in ESS10: Austria, Belgium, Czechia, Finland, France, Hungary, Iceland, Italy, Portugal, Slovenia, Sweden and the United Kingdom – as a follow-up panel survey across six waves [13]. Since the data utilised in this study was taken from wave 4 (when mental health was assessed), this limited the sample to 11 countries (excluding Hungary). The CRONOS-2 web panel included additional inclusion criteria; participants were required to be 18 years or older and needed internet access [13]. Since participants were recruited after taking part in ESS10, this allowed for panel member recruitment from random probability samples and longitudinal follow-up. The two datasets (ESS10 and CRONOS-2 wave 4) were merged together using the participant identification number, country and survey mode. A total of 18,637 ESS10 respondents were eligible for CRONOS-2 recruitment [13].

The two surveys were conducted in accordance to the Declaration of Professional Ethics of the International Statistical Institute [14]. Ethical approval was not required for this secondary data analysis. The article complies with the STrengthening the Reporting of OBservational studies in Epidemiology (STROBE) guidelines [15].

### Exposure, outcomes and covariates

Climate change worry (exposure variable) was measured via the question which asked, “How worried are you about climate change?”, during the ESS10 survey (baseline). Respondents were asked to choose an option from a 5-point Likert scale which ranged from 1 (not at all worried) to 5 (extremely worried). The responses “not at all worried”, “not very worried” and “somewhat worried” were grouped as “not worried about climate change”, whereas the responses “very worried” and “extremely worried” were grouped as “worried about climate change” to form a binary variable.

To measure depression (outcome variable), the Center for Epidemiologic Studies - Depression Scale (CES-D 8) [16] block of questions was utilised from CRONOS-2 wave 4. These questions form an eight-item adapted version of the original CES-D questionnaire [17] – a 20 item measure asking respondents to rate how often over the past week they have experienced symptoms of depression. The four possible response options were “rarely or none of the time”, “some of the time”, “often” and “most of the time”, which were scored from 0 to 3, respectively. To assess anxiety (outcome variable), the Generalised Anxiety Disorder Assessment (GAD-7) block of questions [18] was utilised from the CRONOS-2 wave 4 web panel. These questions explore how often respondents experience symptoms of anxiety over the past two weeks. The four possible response options were “rarely or none of the time”, “some of the time”, “often” and “most of the time”. The responses were scored from 0 to 3 respectively. For depression and anxiety, the overall scores for each block of questions were calculated, and a cut-off of 10 was applied for potential clinical (moderate/severe) depression or anxiety, according to relevant guidelines [18], [19].

To assess sleep disturbance (outcome variable), one of the questions from the CES-D 8 block was utilised, which asked respondents how often in the past week they had experienced restless sleep. Sleep disturbance was converted into a binary variable by grouping the responses “rarely or none of the time” and “some of the time” as not experiencing sleep disturbance, and grouping “often” and “most or all of the time” as positive for experiencing sleep disturbance.

Baseline characteristic data of the respondents was obtained from the ESS10 survey. This included the age of respondents, gender, education level, income level, whether they lived in a rural or urban setting, their country of residence and whether they were a migrant. These were included as model covariates as they were thought to potentially confound the relationship between climate change worry and the outcomes. Education level was determined in ESS10 using an adapted version of the International Standard Classification of Education (ISCED) 2011 [20]. The European Survey-ISCED (ES-ISCED) split highest educational attainment into 8 tiers, ranging from less than primary education to a doctoral degree or equivalent [21]. This was categorised into low, intermediate and high levels of education by grouping ES-ISCED 1 to 2, ES-ISCED 3 to 5, and ES-ISCED 6 to 8 respectively.

A decile approach was utilised to measure income, based on national deciles of the actual household income range in each country [21]. Respondents estimated their household’s total income from all sources, after tax and compulsory deductions, and selected one of 10 income deciles which applied to them. Income levels were then grouped into low, middle and high income by grouping deciles 1 to 3, deciles 4 to 7 and deciles 8 to 10, respectively.

To assess living in an urban or rural setting, respondents chose the most appropriate description of their domicile, which ranged from “a big city”, “suburbs or outskirts of a big city”, “town or small city”, “country village” to “farm or home in countryside” [21].

The former three categories were grouped as urban and the latter two were grouped as rural. Migration status was derived from a survey item which asked participants whether they were born in their current country of residence (yes/no).

### Sources of bias

Post-stratification weights for CRONOS-2 developed by the ESS team were applied during the analysis to minimise the impact of non-response bias and attrition. This incorporated the post-stratified ESS10 design weight and adjustments for non-response at both ESS10 and the respective CRONOS-2 wave [13].

### Statistical analysis

Three analyses were conducted to investigate the research questions. First, to determine which countries had the highest levels of climate change worry, climate change worry was summarised descriptively using percentages and graphs, stratifying respondents by country. Next, to investigate the relationship between climate change worry and risk of depression, anxiety and sleep disturbance, two logistic regression analyses were applied. The associations were reported as odds ratios (OR) with 95% confidence intervals (CI).

The first set of models investigated the relationship between climate change worry and depression, anxiety and sleep disturbance in a pooled cross-country analysis. Great Britain was chosen as the reference country for all analyses. The first model of this analysis adjusted for country fixed effects, the second model adjusted for country fixed effects, age and gender, and the third model adjusted for country, age, gender, migrant status, urban/rural status, education and income level. The second set of models investigated the relationship between climate change worry and depression, anxiety and sleep disturbance whilst stratifying by country to investigate potential differences between countries, adjusting for confounders as in the first set of models.

Respondents who had missing values for any of the variables included in the models were excluded from the analyses to conduct a complete case analysis. All tables and analyses were weighted as recommended [13]. All analyses were performed using Stata/MP 18.0.

## FINDINGS

A total of 6,032 of the eligible respondents took part in CRONOS-2 Wave 4, and 877 (14.5%) respondents were excluded due to missing data, limiting the number of respondents included in this study to 5,155 across the 11 countries. Most of the missing data stemmed from baseline income level (493 missing values) and mental health in the CRONOS-2 wave 4 questionnaire (386 missing values). The baseline sociodemographic characteristics of the respondents for each country are summarised in Table 1.

**Table 1.** Baseline characteristics of the 5,155 respondents who participated in ESS10 and CRONOS-2 wave 4*.

| Country | Sample size | Mean Age (SD) | Gender |  | Income level |  |  | Education level |  |  | Urban or rural residency |  | Migrant |  |
| --- | --- | --- | --- | --- | --- | --- | --- | --- | --- | --- | --- | --- | --- | --- |
|  |  |  | male (%) | female (%) | low (%) | middle (%) | high (%) | low (%) | intermediate (%) | high (%) | rural (%) | urban (%) | no (%) | yes (%) |
| **Austria** | 537 | 47 (17.9) | 56.4 | 43.6 | 14.2 | 39.1 | 46.7 | 11.3 | 60.4 | 28.2 | 38.9 | 61.1 | 85.4 | 14.6 |
| **Belgium** | 501 | 48.5 (16.4) | 52.7 | 47.4 | 14.5 | 50.4 | 35.2 | 18.9 | 37.9 | 43.2 | 55.4 | 41.1 | 83.2 | 16.8 |
| **Czechia** | 180 | 52.2 (15.3) | 40.8 | 59.2 | 24.6 | 55.4 | 20.0 | 6.9 | 76.5 | 16.7 | 20.5 | 79.5 | 97.0 | 3.0 |
| **Finland** | 697 | 50.4 (16.9) | 46.1 | 53.9 | 16.7 | 45.5 | 37.7 | 16.7 | 47.2 | 36.1 | 30.1 | 69.9 | 96.5 | 3.5 |
| **France** | 641 | 47 (16.3) | 45.4 | 54.6 | 16.6 | 40.3 | 43.1 | 15.9 | 59.2 | 25.0 | 39.0 | 61.0 | 89.1 | 10.9 |
| **Great Britain** | 431 | 50.2 (16.7) | 47.8 | 52.2 | 20.3 | 41.6 | 38.0 | 18.9 | 38.7 | 42.4 | 26.0 | 74.0 | 82.9 | 17.1 |
| **Iceland** | 472 | 47 (16.7) | 49.9 | 50.1 | 23.3 | 54.8 | 22.0 | 24.0 | 42.5 | 33.5 | 15.7 | 84.3 | 92.1 | 7.9 |
| **Italy** | 153 | 42.1 (17.2) | 50.7 | 49.3 | 21.9 | 55.3 | 22.8 | 33.7 | 47.9 | 18.4 | 49.2 | 50.8 | 93.3 | 6.7 |
| **Portugal** | 275 | 45.7 (14.9) | 45.7 | 54.3 | 13.8 | 55.3 | 31.0 | 40.3 | 32.7 | 26.9 | 26.1 | 73.9 | 85.9 | 14.1 |
| **Sweden** | 779 | 47.6 (17.6) | 52.1 | 47.9 | 13.6 | 49.7 | 36.7 | 15.9 | 51.6 | 32.5 | 23.8 | 76.2 | 86.3 | 13.7 |
| **Slovenia** | 489 | 44.7 (15.3) | 50.5 | 49.5 | 18.3 | 52.0 | 29.7 | 6.7 | 58.7 | 34.6 | 51.8 | 48.2 | 88.4 | 11.6 |

\*Weighted. SD=standard deviation

Levels of climate change worry by country are illustrated in Figure 1. The level of climate worry was highest in Portugal (63.7%), followed by Austria (53.2%) and Slovenia (51.3%), and lowest in Italy (30.2%), followed by Sweden (31.6%) and Iceland (32.5%).

**Figure 1.**
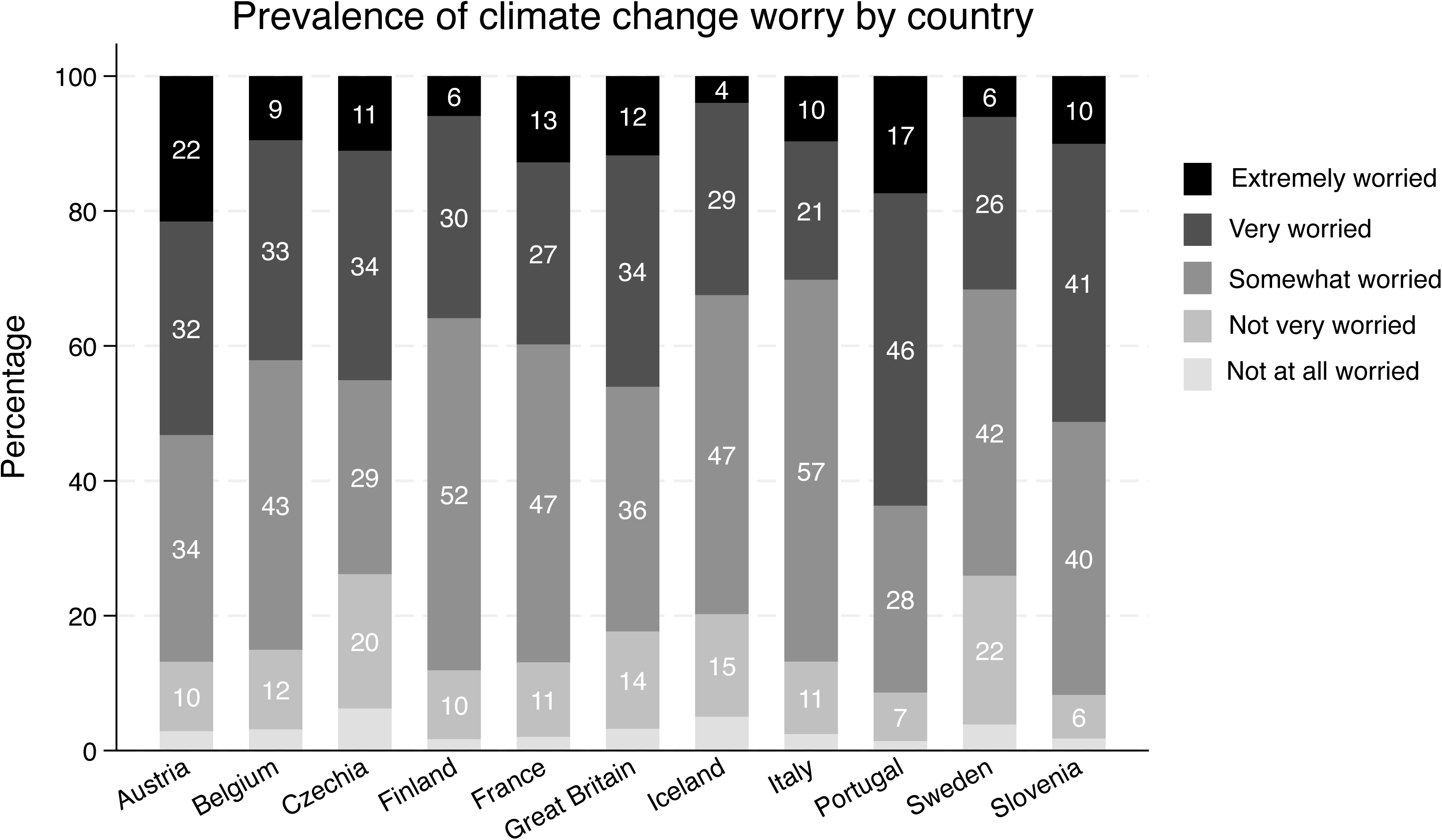

Results of the first set of logistic regression models showing the adjusted relationship between worry about climate change and depression, anxiety and sleep disturbance are summarised in Table 2. Worry about climate change was associated with increased risk of anxiety (OR: 1.38, 95% CI: 1.13, 1.68), but not depression (OR: 1.10, 95% CI: 0.94, 1.29) or sleep disturbance (OR: 1.08, 95% CI: 0.92, 1.27). A decreased risk of depression, anxiety and sleep disturbance was generally associated with higher education and income groups. Women had higher risk of depression, anxiety and sleep disturbance compared to men, whilst greater age was associated with a decreased risk in depression, anxiety and sleep disturbance. In terms of countries, living in the Czech Republic was associated with increased risk of depression and anxiety compared to Great Britain, but not sleep disturbance. On the other hand, living in Finland was associated with a decreased risk for all three outcomes, compared to Great Britain.

**Table 2.** Results from models assessing risk of depression, anxiety and sleep disturbance according to worry about climate change across 11 countries in the ESS (n = 5,155)

| Variable | Outcome |  |  |
| --- | --- | --- | --- |
|  | Odds ratio (95% CI) |  |  |
|  | Depression | Anxiety | Sleep disturbance |
| Worried about climate change | 1.10<br>(0.94 - 1.29) | 1.38***<br>(1.13 - 1.68) | 1.08<br>(0.92 - 1.27) |
| Austria<sup>†</sup> | 0.65**<br>(0.44 - 0.95) | 0.73<br>(0.44 - 1.20) | 0.46***<br>(0.31 - 0.67) |
| Belgium<sup>†</sup> | 0.83<br>(0.58 - 1.20) | 0.98<br>(0.62 - 1.55) | 0.61***<br>(0.43 - 0.87) |
| Czech Republic<sup>†</sup> | 1.60**<br>(1.01 - 2.52) | 1.84**<br>(1.02 - 3.32) | 0.90<br>(0.57 - 1.41) |
| Finland<sup>†</sup> | 0.45***<br>(0.31 - 0.64) | 0.38***<br>(0.23 - 0.63) | 0.27***<br>(0.19 - 0.38) |
| France<sup>†</sup> | 0.83<br>(0.60 - 1.16) | 1.17<br>(0.77 - 1.78) | 0.75*<br>(0.55 - 1.02) |
| Iceland<sup>†</sup> | 0.56***<br>(0.39 - 0.80) | 0.73<br>(0.45 - 1.19) | 0.36***<br>(0.25 - 0.53) |
| Italy<sup>†</sup> | 1.41<br>(0.88 - 2.27) | 1.72*<br>(0.95 - 3.10) | 0.34***<br>(0.19 - 0.59) |
| Portugal<sup>†</sup> | 1.07<br>(0.73 - 1.57) | 1.24<br>(0.76 - 2.02) | 0.54***<br>(0.37 - 0.81) |
| Sweden<sup>†</sup> | 0.78<br>(0.56 - 1.08) | 0.82<br>(0.52 - 1.27) | 0.43***<br>(0.31 - 0.60) |
| Slovenia<sup>†</sup> | 0.58***<br>(0.41 - 0.83) | 1.06<br>(0.68 - 1.64) | 0.46***<br>(0.33 - 0.65) |
| Age of respondent | 0.97***<br>(0.97 - 0.98) | 0.96***<br>(0.96 - 0.97) | 0.98***<br>(0.98 - 0.99) |
| Female (ref=male) | 1.17*<br>(1.00 - 1.37) | 1.42***<br>(1.16 - 1.75) | 1.30***<br>(1.11 - 1.52) |
| Urban residency (ref=rural) | 1.08<br>(0.91 - 1.29) | 1.14<br>(0.92 - 1.42) | 1.00<br>(0.84 - 1.19) |
| Intermediate level education<sup>‡</sup> | 0.61***<br>(0.48 - 0.78) | 0.69**<br>(0.50 - 0.93) | 0.81<br>(0.62 - 1.05) |
| High level education<sup>‡</sup> | 0.44***<br>(0.34 - 0.57) | 0.47***<br>(0.34 - 0.66) | 0.70**<br>(0.53 - 0.92) |
| Middle-income<sup>§</sup> | 0.56***<br>(0.46 - 0.69) | 0.65***<br>(0.50 - 0.83) | 0.79**<br>(0.64 - 0.97) |
| High-income<sup>§</sup> | 0.38***<br>(0.30 - 0.48) | 0.53***<br>(0.39 - 0.70) | 0.65***<br>(0.51 - 0.82) |
| Migrant (ref=non-migrant) | 0.90<br>(0.68 - 1.20) | 0.99<br>(0.70 - 1.38) | 0.87<br>(0.65 - 1.16) |

\*\*\* p<0.01, \*\* p<0.05, \* p<0.1
‡ reference is low level education
† reference is Great Britain
§ reference is low-income

The risk of depression, anxiety and sleep disturbance according to worry about climate change in each country is summarised in Table 3. Worry about climate change was most strongly associated with depression in Austria (OR: 1.82, 95% CI: 1.02, 3.25), Iceland (OR: 1.71, 1.00, 2.93) and Belgium (OR: 1.69, 95% CI: 1.04, 2.75), with anxiety in Italy (OR: 2.82, 95% CI: 1.00, 7.95) and Slovenia (OR: 2.00, 95% CI: 1.18, 3.39), and with sleep disturbance in Belgium (OR: 1.93, 95% CI: 1.19, 3.13).

**Table 3.** Stratified results for each of the 11 countries in the ESS 10 dervied from models assessing risk of depression, anxiety and sleep disturbance according to worry about climate change (n=5,155).

| Country | Outcome |  |  |
| --- | --- | --- | --- |
|  | Odds ratio (95% CI) |  |  |
|  | Depression | Anxiety | Sleep disturbance |
| Austria | 1.82** (1.02 - 3.25) | 1.34 (0.71 - 2.55) | 0.91 (0.52 - 1.57) |
| Belgium | 1.69** (1.04 - 2.76) | 1.58 (0.87 - 2.89) | 1.93*** (1.19 - 3.13) |
| Czech Republic | 0.92 (0.43 - 1.95) | 1.23 (0.48 - 3.10) | 1.10 (0.50 - 2.43) |
| Finland | 0.97 (0.58 - 1.64) | 1.14 (0.53 - 2.45) | 1.10 (0.66 - 1.83) |
| France | 1.00 (0.67 - 1.49) | 1.42 (0.88 - 2.28) | 1.07 (0.74 - 1.56) |
| Great Britain | 0.73 (0.43 - 1.23) | 1.16 (0.57 - 2.37) | 0.66 (0.40 - 1.10) |
| Iceland | 1.71* (1.00 - 2.93) | 1.22 (0.58 - 2.57) | 0.78 (0.41 - 1.48) |
| Italy | 1.66 (0.59 - 4.72) | 2.82** (1.00 - 7.95) | 1.50 (0.60 - 3.76) |
| Portugal | 0.85 (0.45 - 1.58) | 1.03 (0.48 - 2.22) | 1.90* (0.91 - 3.98) |
| Sweden | 0.72 (0.47 - 1.11) | 1.27 (0.73 - 2.24) | 0.85 (0.56 - 1.29) |
| Slovenia | 1.39 (0.87 - 2.20) | 2.00** (1.18 - 3.39) | 1.41 (0.91 - 2.19) |

\*\*\* p<0.01, \*\* p<0.05, \* p<0.1
Models adjusted for age, gender, education level, income level, urban/rural residency and migrant status

## DISCUSSION

Our study assessed levels of climate change worry across Europe and its relationship with depression, anxiety and sleep disturbance. The highest levels of climate change worry were found in Portugal, followed by Austria and Slovenia.

Associations between climate change worry and at least one outcome were found in Austria and Slovenia, but not Portugal. Overall, high levels of climate change worry were associated with an increased risk of clinical levels of anxiety in the pooled analyses across all countries. When stratifying the models by country, the estimates were stronger for some countries. There was an increased risk of depression by around 9.8% and 10.1% in Austria and Belgium, respectively. The increase in risk of anxiety was around 15.4% in Italy and 9.3% in Slovenia, although Italy’s results should be interpreted with caution due to having a smaller sample size. In Belgium, there was an associated 12.6% increase in risk of sleep disturbance among people with high levels of climate worry.

Importantly, although not statistically significant, some findings for particular countries suggested that climate change worry may be a protective factor against depression and sleep disturbance. WHO describes the “dual impact of involvement in climate change action”, whereby engaging in pro-environmental behaviours could bring about potentially beneficial mental health outcomes [2]. This is of course in addition to the possibly stressful experience of taking up climate action.

Nevertheless, this could account for the potentially protective role of climate change worry against depression in countries like Great Britain and Sweden.

Overall, the results imply that in parts of Europe, climate change worry is not strongly associated with mental health. However, this is not homogenous, with the population in some countries being more worried about climate change, which could be affecting mental health, particularly levels of anxiety. This is possibly because some countries are at higher risk of the direct effects of climate change, such as heatwaves and wildfire. For instance, increased risk of drought and forest fires in the south of Europe may have led to higher levels of climate change worry in Portugal [22]. While all countries Europe will be affected by climate change in some way, the possible impacts in other areas are perhaps less severe.

### Strengths and limitations of this study

Strengths of this study include its design and cross-national approach. This allowed for temporal assessment of the exposure and outcomes in a more representative sample across Europe, increasing its external validity in comparison to existing research. In addition, this study examined three important outcomes using validated measures, allowing for further understanding in the interplay of depression, anxiety and sleep disturbance with climate change worry.

However, there were also several limitations. The number of respondents from each country was inconsistent, with smaller samples in some countries limiting the precision of model estimates. Respondents in this study had to be 18 years or older to take part in CRONOS-2. Previous studies have indicated that youth and children are more likely to be aware of climate change impacts and are more vulnerable to these impacts [3][5]. As a result, the overall levels of climate change worry and its impact on mental health may be underestimated in this study. A significant portion of the missing values stemmed from the CRONOS-2 online panel from the mental health questionnaire. This potentially introduces bias, particularly for the Czech Republic and Portugal whereby 17% and 9% of respondents had to be excluded (respectively) due to missing information regarding mental health. One previous study suggests underreporting of mental disorders due to stigmatisation and association with negative stereotypes [23], indicating that individuals with mental disorders are perhaps less likely to answer mental health related questions.

CRONOS-2 was conducted via an online survey, therefore excluding people without internet access which may introduce some bias. Furthermore, using differing methods of data collection may introduce variability into findings [24], as well as variability in response rates [25]. Lastly, despite the findings of this study, the study design cannot attribute poor mental health to climate change worry. Further research with repeated measures of climate change worry and mental health outcomes is required to establish causality.

## CLINICAL IMPLICATIONS

Further research exploring reasons for differences in findings between countries is required, looking into factors, such as government policies and risk of extreme weather events within specific countries. Another gap urgently needing addressed is the lack of practice frameworks and policies to help practitioners support individuals presenting with poor mental health driven by climate change worry and anxiety. This phenomenon does not fit traditional mental illness models and is unique in the sense that individuals may experience distressing and debilitating emotions without necessarily going through the traumatic direct effects of climate change. Standard treatment for mental disorders were not designed to treat real and imminent threats. Therefore, research into development of robust and holistic approaches to overcome mental health issues due to climate anxiety is imperative [3].

## Data Availability

All data used are openly available (upon registration) at https://www.europeansocialsurvey.org/.

## Acknowledgements

We thank the participants of the ESS and the ESS team for supplying the data. This research was previously presented at the 2024 Society for Social Medicine and Population Health Annual Scientific Meeting and the conference abstract is published in the Journal of Epidemiology and Community Health.

## Author contributorship

AC: writing—original draft (lead), conceptualisation (supporting), methodology (supporting), formal analysis (lead). CLN: writing—reviewing and editing (supporting), methodology (lead), conceptualisation (lead), data curation (lead), supervision (lead), formal analysis (supporting), guarantor.

## Data sharing

ESS data are openly available (upon registration) at https://www.europeansocialsurvey.org/.

## Patient consent for publication

Not applicable.

## Ethical approval

Ethical approval was not required for this secondary data analysis. Details of the ESS research ethics processes can be found here: https://www.europeansocialsurvey.org/about/research-ethics.

